# The Association Between Gut-Derived Metabolites, Body Composition, and Energy Expenditures in Adolescents: A Sex-Stratified Analysis of the COPSAC2000 Study

**DOI:** 10.64898/2026.02.11.26346082

**Authors:** Negar Chahibakhsh, David Horner, Bo Chawes, Rebecca Vinding, Ann-Marie M. Schoos, Zhuobing Peng, Shiraz Shah, Parvaneh Ebrahimi, Morten A. Rasmussen

## Abstract

The gut microbiota has been implicated in regulating body composition, insulin resistance, and energy metabolism through microbial metabolites, including short-chain fatty acids (SCFAs) and amino acids. However, evidence in adolescents, particularly regarding sex-specific differences and lifestyle such as alcohol intake, remains limited. Characterizing sex-specific metabolic signatures in adolescence may improve early identification of metabolic risk.

To address this gap, we investigated associations between fecal metabolites, body composition, insulin resistance, and energy expenditure in 158 adolescents aged 18 from the Copenhagen Prospective Studies on Asthma in Childhood (COPSAC2000).

Quantitative fecal metabolomics was performed using proton nuclear magnetic resonance (^1^H-NMR) spectroscopy, profiling 32 metabolites. Associations with body composition, insulin resistance, and energy expenditure were evaluated using sex-stratified univariate and multivariate modelling with false discovery rate (FDR ≤ 0.05 and 0.2).

Fecal acetate and ethanol were more associated with fat-free mass index (FFMI) and waist-to-height ratio (WHtR) than with body mass index (BMI) in females; in males, no associations remained after FDR. Lysine and leucine showed associations with BMI and insulin resistance in females. Acetate, butyrate, glucose, and methanol were associated with total energy expenditure (TEE) in females, whereas no association survived in males. Alcohol intake was positively associated with fecal ethanol, glucose, and methanol, and inversely with trimethylamine in females, while galactose showed a positive association in males.

These findings demonstrate that gut microbiota–derived metabolites are related to body composition, insulin sensitivity, and energy balance in adolescents, particularly females, highlighting the utility of fecal metabolomics in exploring mechanisms underlying metabolic variation.

## 1. Introduction

The human gut microbiota is increasingly recognized as a dynamic contributor to host metabolic health, both through interactions with the epithelial barrier (Thaiss et al., 2016) and via metabolic products (Koh et al., 2016). Through the fermentation of dietary fibers and proteins, gut microbes generate metabolites, including SCFAs and amino acid–derived compounds, which influence nutrient metabolism, energy balance, body composition, and modulate host physiology (Hong et al., 2025; Lange et al., 2023).

Fecal metabolomics has emerged as a non-invasive approach to study host-microbiome interactions, providing insights into microbial activity and its potential influence on systemic metabolism and health outcomes (Zierer et al., 2018). Altered fecal metabolite profiles have been associated with obesity, insulin resistance, and differences in body composition (Visconti et al., 2019). For instance, obese individuals often exhibit distinct SCFAs and amino acids compared to lean individuals (Hong et al., 2025).

A key unresolved question is when, during the life course, gut microbial signals become detectably linked to adiposity and body composition. The early-life programming hypothesis proposes that obesity-related microbial features are established in infancy with long-term metabolic consequences. However, recent longitudinal evidence from a deeply phenotyped COPSAC2000 birth cohort reported no consistent associations between early-life gut microbiota composition and later childhood adiposity. This suggests that gut-related metabolic signals may not act as stable early-life predictors, but may emerge later as functional outputs influenced by maturation and lifestyle exposures (Yan et al., 2024).

Recent studies also suggest that gut microbiota and their metabolites can influence basal metabolic rate (BMR) and total energy expenditure (TEE), possibly by modulating mitochondrial efficiency, substrate utilization, and appetite-regulating hormones (Chevalier et al., 2015; Clarke et al., 2014). However, evidence in adolescents remains limited, as this period is characterized by ongoing metabolic and developmental transitions.

Also, lifestyle factors, including alcohol consumption, may contribute to inter-individual variability in the gut metabolome. Alcohol intake can alter gut microbiota composition, intestinal permeability, and microbial metabolic pathways, with potential implications for energy regulation and adiposity (Wang et al., 2023). Emerging human data suggest that these changes may be reflected in fecal metabolomic signatures (Zierer et al., 2018).

Despite growing interest in the gut-metabolism axis, few studies have comprehensively examined the associations between fecal metabolites and metabolic health markers, including body composition and energy expenditure, in general adolescent populations. Furthermore, the role of sex differences and the impact of alcohol consumption on gut-derived metabolites in youth remains poorly understood. Evidence from randomized trials shows that lifestyle factors preferentially modify body composition-sensitive traits, such as fat mass and waist-related measures, rather than BMI alone, supporting the use of metabolomics as a sensitive readout of lifestyle-induced metabolic variation (Chahibakhsh et al., 2024).

We hypothesized that gut-derived metabolites are associated with body composition, energy expenditure, and insulin resistance, with potential sex differences and modification by alcohol intake. To address these gaps, we investigated fecal metabolite profiles, including SCFAs, amino acids, and related compounds, in adolescents from the COPSAC2000 study. We explored associations between fecal metabolite composition and: 1) body composition, particularly BMI, FFMI, WHtR, and insulin resistance, 2) alcohol consumption, and 3) energy expenditure, specifically BMR and TEE, in 18-year-old adolescents.

## 2. Methods

### 2.1. Study population and variables

Participants were part of COPSAC2000, a prospective clinical mother-child cohort study of 411 children of asthmatic mothers followed longitudinally since age one month (Bisgaard, 2004; Shojaeifard et al., 2024). At age 18, fecal samples were collected from 185 participants, of whom 27 subjects were excluded due to missing anthropometric data, resulting in 158 participants (89 females, 69 males).

32 fecal metabolites were analyzed, grouped into six biological categories: amino acids, SCFAs, carbohydrates/sugars, alcohols and related compounds, energy metabolism intermediates, and nitrogenous compounds.

### 2.2. Metabolic profiling and extracting metabolic features

Fecal metabolites were measured using a high-throughput nuclear magnetic resonance ¹H-NMR platform for 185 fecal samples from the subjects, with absolute quantification performed within the SigMa framework using an electronic reference-based approach (Cui et al., 2021). Blank control samples were analyzed alongside fecal samples to monitor background signals and potential contamination. Glycerol was added to fecal samples before freezing to preserve sample integrity during storage, and glycerol-derived signals were excluded before data analysis. No ethanol was added during storage.

### 2.3. Body anthropometrics and basal metabolism rate

Anthropometrics were assessed at the 18-year visit. Weight was measured using calibrated digital scales (barefoot and with lightweight clothes), and height using a stadiometer, calibrated annually. Hip circumference was measured at the widest girth around the hip (cm), and the waist circumference was measured using a tape, placing it around the navel as a reference point and averaging two measurements taken during inspiration and expiration. WHtR was calculated by dividing the waist circumference (cm) by the height (cm), and WHR was determined by dividing the waist circumference (cm) by the hip circumference (cm) (Chahibakhsh et al., 2019).

The Bioelectrical Impedance Analysis (BIA) was applied (TANITA MC-780MA) to assess body composition, including skeletal muscle mass, fat percentage, and distribution, BMI, fat-free mass (FFM), and indexes, including FFMI and FMI. The measurements were conducted following standardized protocols (Shojaeifard et al., 2024; Yan et al., 2024).

### 2.4. Insulin resistance

The homeostasis model assessment of insulin resistance (HOMA-IR) index was used to estimate insulin resistance, based on the fasting blood glucose and insulin levels(Chahibakhsh et al., 2019; Matthews et al., 1985). Fasting blood glucose was measured by enzymatic determination and absorption photometry (Siemens Atellica CH 930), and fasting insulin was measured using sandwich immunoassays with chemiluminescence (Siemens Atellica IM).

### 2.5. Physical activity, BMR, and TEE

Physical activity was assessed spontaneously using an omnidirectional accelerometer (ActiGraph wGT3X+ActiLife software, ActiGraph LLC, Pensacola, FL, USA) worn on the non-dominant wrist for 2 weeks to capture both daytime activity and sleep. Accelerometer data were processed using ActiLife software to derive time spent at different activity intensities (min/day). According to these measures, Physical Activity Levels (PAL) were classified as sedentary (< 150 min/day), lightly active (150-300 min/day), moderately active (300-450 min/day), or very active (> 450 min/day).

BMR was estimated using the Mifflin–St. Jeor’s equation (Mahan, 2016), a validated and widely used method in population-based studies:

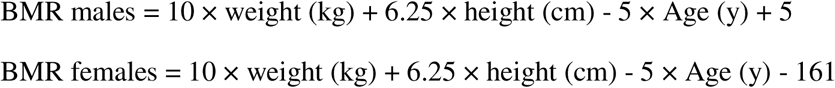

TEE was estimated using a 24-hour time-weighted metabolic equivalent (MET) approach. Time spent in each activity category (sleep, sedentary, light, moderate, and vigorous) was summed and normalized to 24 hours, and TEE was calculated as:

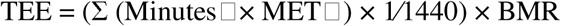

where MinutesC represents time spent in each activity, and METC the corresponding intensity of that category.

Literature-based MET values were applied as time-weighted coefficients for each activity: sleep (0.95), sedentary (1.3), light (2.3), moderate (4.5), and vigorous (8.0), consistent with standard MET classification from Compendium of Physical Activities (Herrmann et al., 2024) and supported by experimental estimates for sitting, sleeping, and walking METs (Mansoubi et al., 2015),(Cho et al., 2016).

### 2.6. Alcohol consumption

Alcohol consumption was examined as an exploratory lifestyle factor, motivated by associations observed for alcohol-related fecal metabolites in the multivariate analyses. We assessed a self-reported questionnaire at age 18, which covers five questions: drinking initiation, frequency, quantity, and binge drinking behavior (Supplementary Methods). One unit of alcohol was defined following the Danish Health Authority’s guidelines as 12 grams of pure alcohol (Jensen et al., 2020).

### 2.7. Statistical data analysis

We explored multivariate relationships between stool metabolites and body anthropometric traits, including BMI, FM, FMI, FFMI, muscle fat ratio, WHtR, and WHR, along with estimated BMR and TEE, while accounting for potential collinearity between body composition measures and energy expenditures. False discovery rate (FDR) correction was applied using the Benjamini–Hochberg procedure. For exploratory screening analyses involving several metabolite–outcome tests, an FDR threshold of 20% was used. For energy-related outcomes (BMR and TEE), a more stringent FDR threshold of 5% was applied to ensure robust inference.

All the statistical analyses were performed in the R environment (version 4.4.1). Univariate linear regression models were used to screen for associations between individual fecal metabolites and body composition, energy expenditure, alcohol consumption, and HOMA-IR, followed by multivariate linear regression models including selected metabolites. All models were implemented using the ‘caret’ package.

Before analysis, the metabolic concentration features were centered, scaled, and log10 normalized. To explore the relationships between stool metabolites and body compositions, we used principal component analysis (PCA) and partial least squares (PLS2) regression, performed using the ‘mixomics’, ‘ggplot2’, and ‘pls’ packages. Model performance was assessed using the mean squared error of cross-validation (MSECV) with leave-one-out cross-validation. Given established sex-specific differences in body composition, energy metabolism, and gut microbiota–related metabolic pathways, all statistical analyses were stratified by sex.

## 3. Results

### 3.1. Study population and data overview

We analyzed COPSAC2000 185 stool samples using the SigMa method. ^1^H-NMR quantification yielded absolute concentrations of 32 metabolites across six categories, including amino acids, SCFAs, carbohydrate/sugars, alcohols and related compounds, energy metabolism intermediates, and nitrogenous compounds for both sexes at age 18. The study design is illustrated in **Figure 1**, sex-stratified anthropometrics are summarized in **Table 1**, and baseline sex-specific fecal metabolite concentrations are presented in **Online Resource 1.**

**Figure 1.**
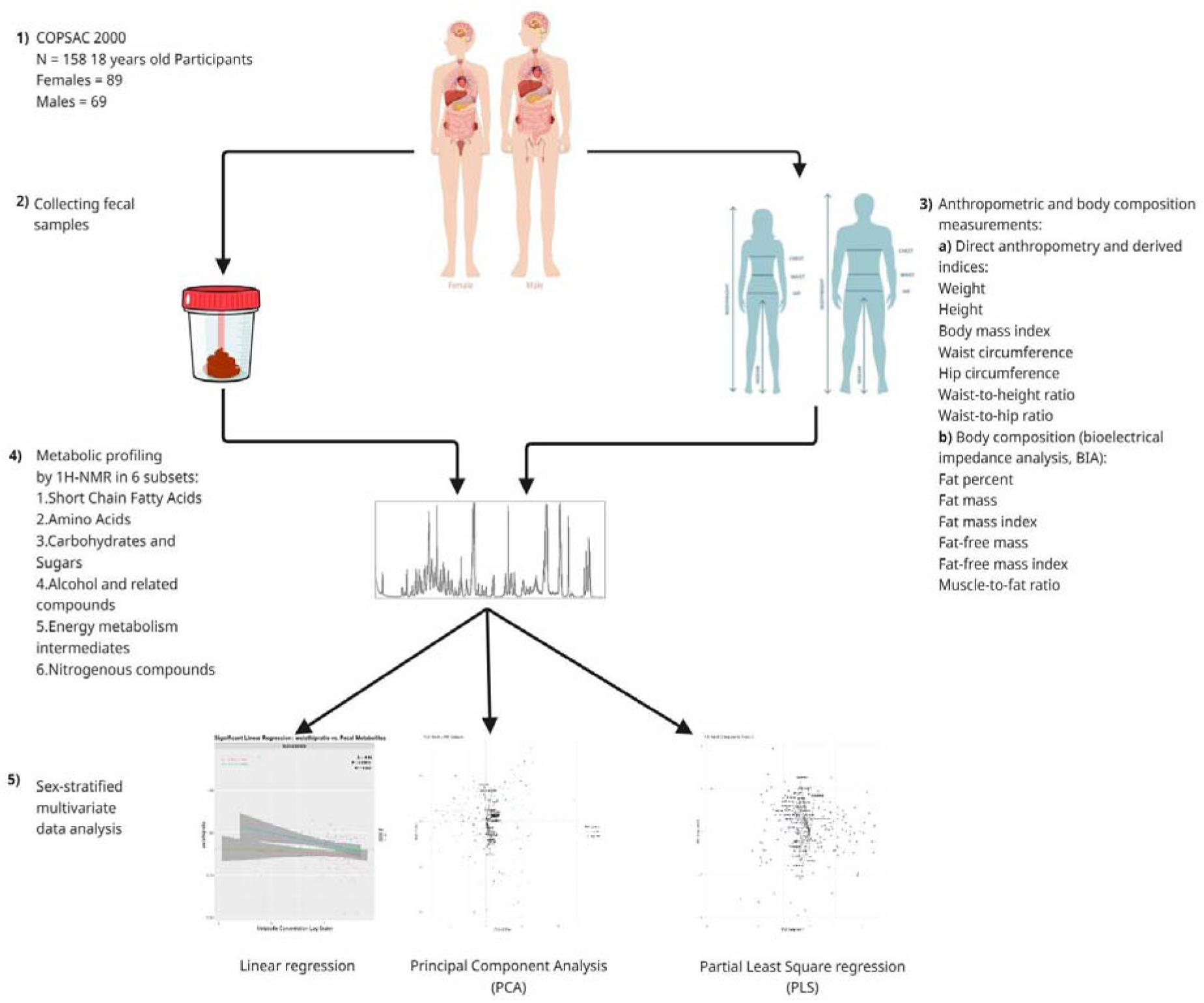
Workflow of the study. Fecal metabolites were profiled using ^1^H-NMR for healthy 18-year-olds from the COPSAC2000 cohort (89 females, 69 males). Anthropometric and derived indices were assessed, alongside body composition parameters obtained by bioelectrical impedance analysis (BIA). Associations between fecal metabolites, anthropometrics, and insulin resistance markers were evaluated using linear regression, PCA, and PLS2 in a sex-stratified manner.

**Table 1.**
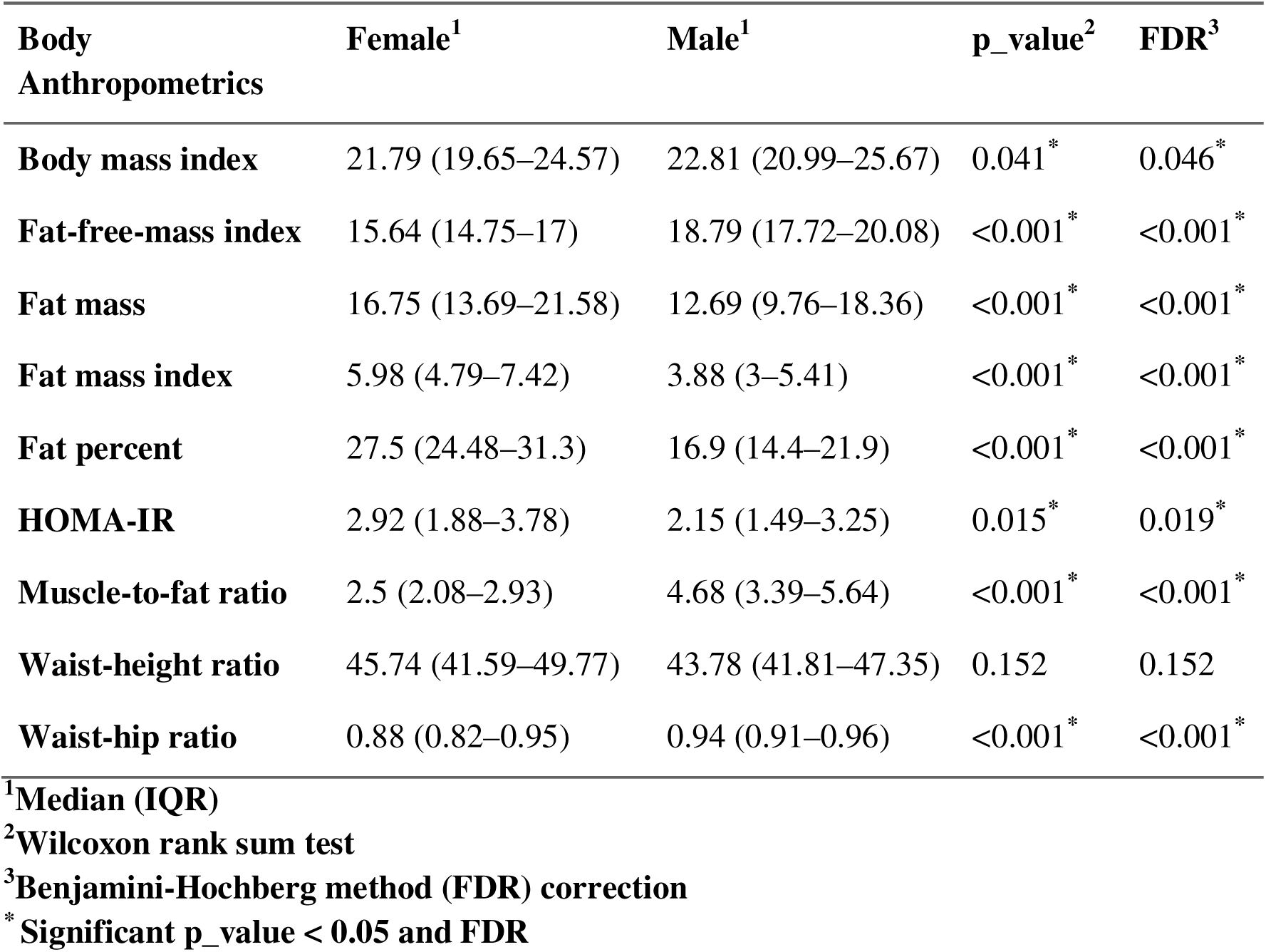
Descriptive anthropometric characteristics of the participants. The first number under each column is the median, and the numbers in the parentheses are the 25th percentile (Q1) and the 75th percentile (Q3), respectively, showing the Inter-Quartile Range (IQR).

### 3.2. Univariate linear regression

Univariate linear regression revealed associations between fecal metabolites and BMI, with significance observed only in females in sex-stratified analyses. Leucine (β = 3.23, p-value =0.047), alanine (β = 3.47, p-value =0.040), lysine (β = 2.91, p-value =0.030), acetate (β = 4. 19, p-value =0.013), propionate (β = 3.09, p-value =0.034) and ethanol (β = 1.77, p-value =0.010) were positively significant. No significant associations were observed in males (p ≥ 0.05). After Benjamini-Hochberg correction (FDR ≤ 0.2), only acetate and ethanol remained significant (adj. R² = 0.03, FDR = 0.198), both for BMI in females (**Figure 2a**).

**Figure 2.**
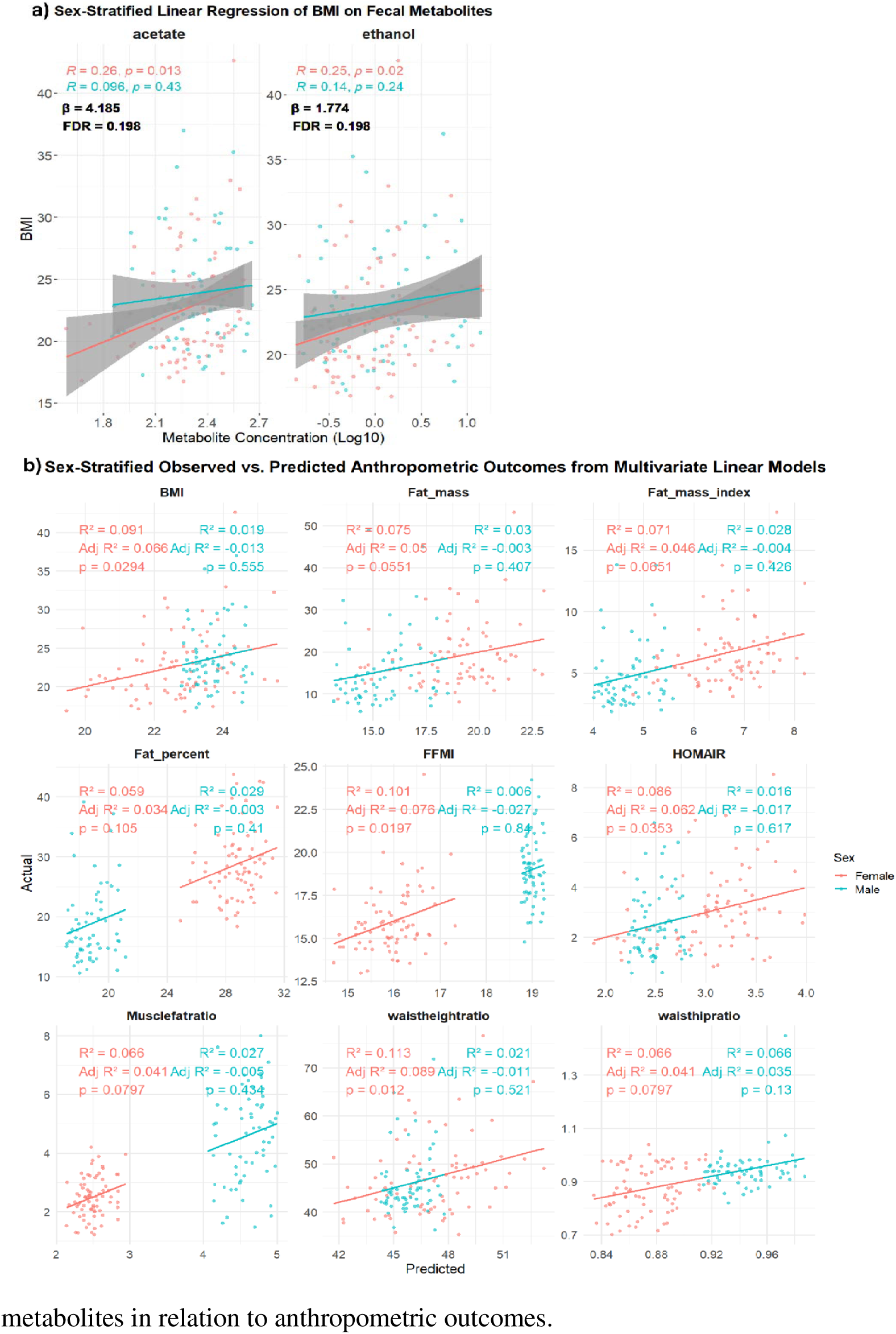
Sex-stratified univariate and multiple linear regression analyses of fecal metabolites in relation to anthropometric outcomes. (a) Sex-stratified linear associations between fecal metabolites and BMI. (b) Sex-stratified observed versus predicted anthropometric outcomes derived from multiple linear regression models.

For FFMI, positive associations were observed for lysine (β = 1.94, p-value =0.012), acetate (β = 1.96, p-value =0.016), and ethanol (β = 0.81, p-value =0.036). Ethanol was also positively associated with WHtR (β = 2.69, p-value =0.022). In sex-stratified analyses, these associations were significant only in females. Although these associations did not remain significant after FDR correction (FDR > 0.2), they showed consistent directional trends across multivariate and PLS2 models (**Figure 4**). No significant associations were observed in males either before or after FDR correction.

**Figure 3.**
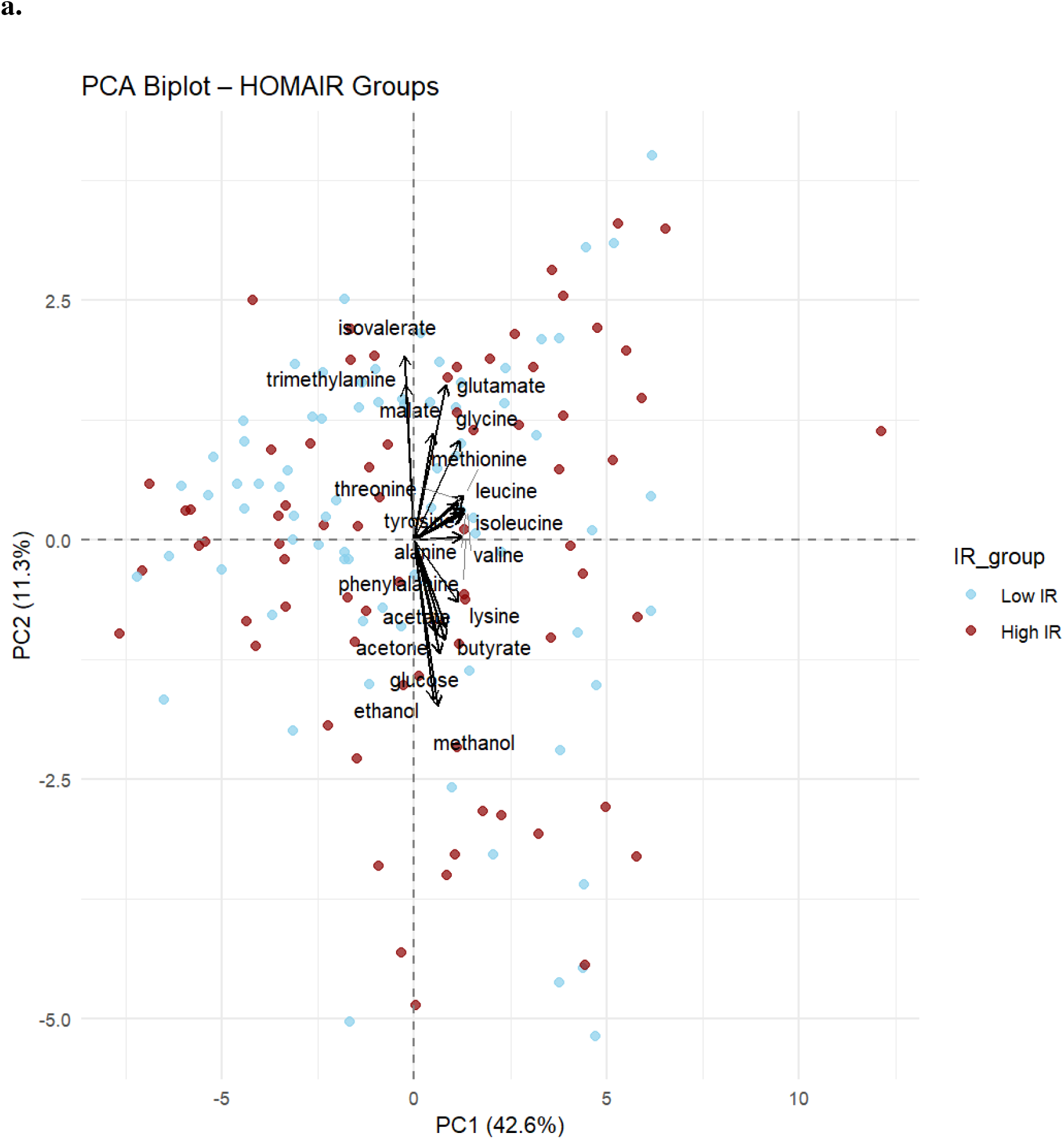
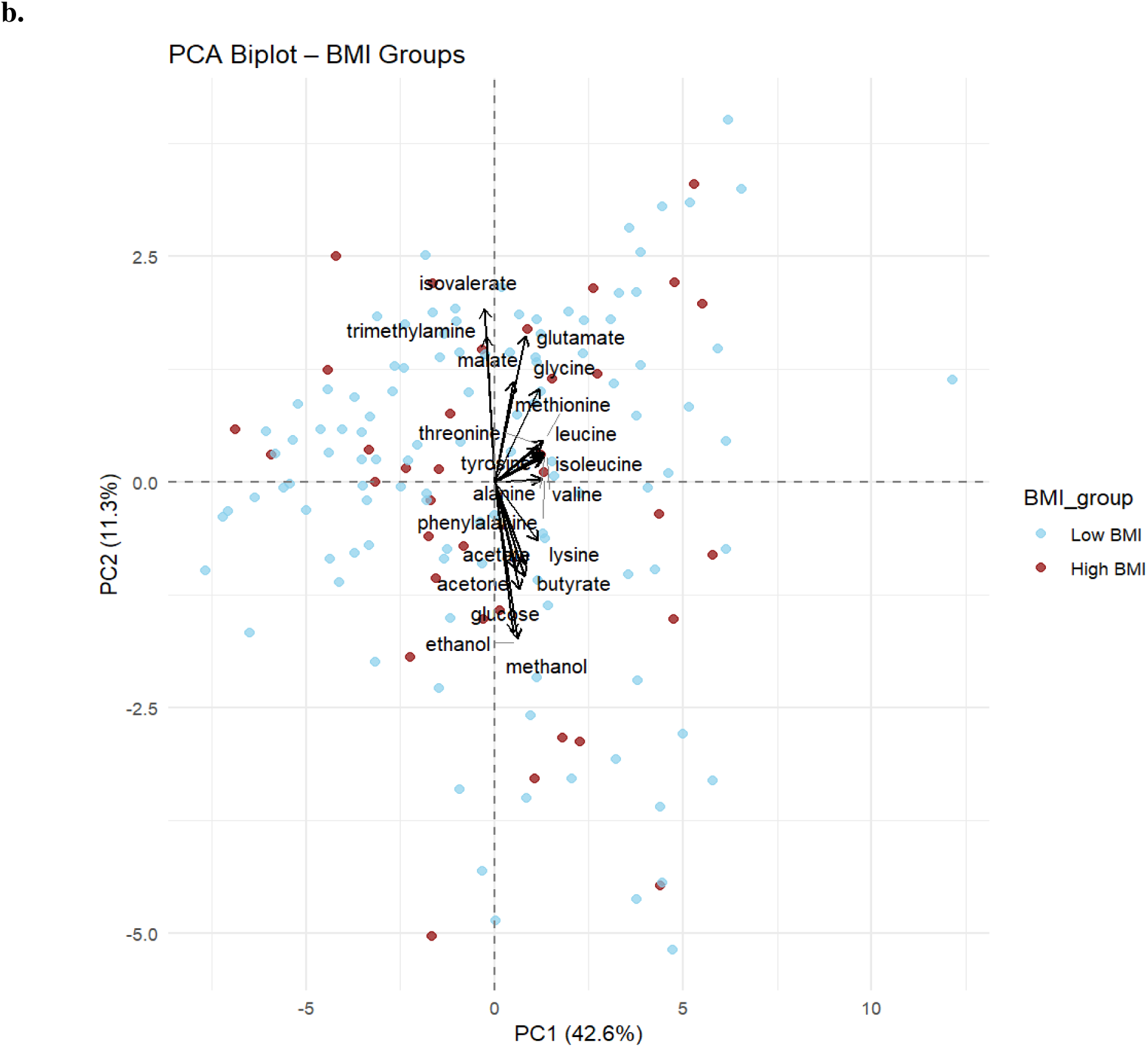
PCA biplots showing PC1 and PC2 for BMI and HOMA-IR stratifications. (a) PCA biplots with distinct visual separation between high and low HOMA-IR groups, (with HOMA-IR > 2.5 as threshold). (b) PCA biplots with distinct visual separation between high and low BMI groups (with BMI > 25 as threshold).

**Figure 4.**
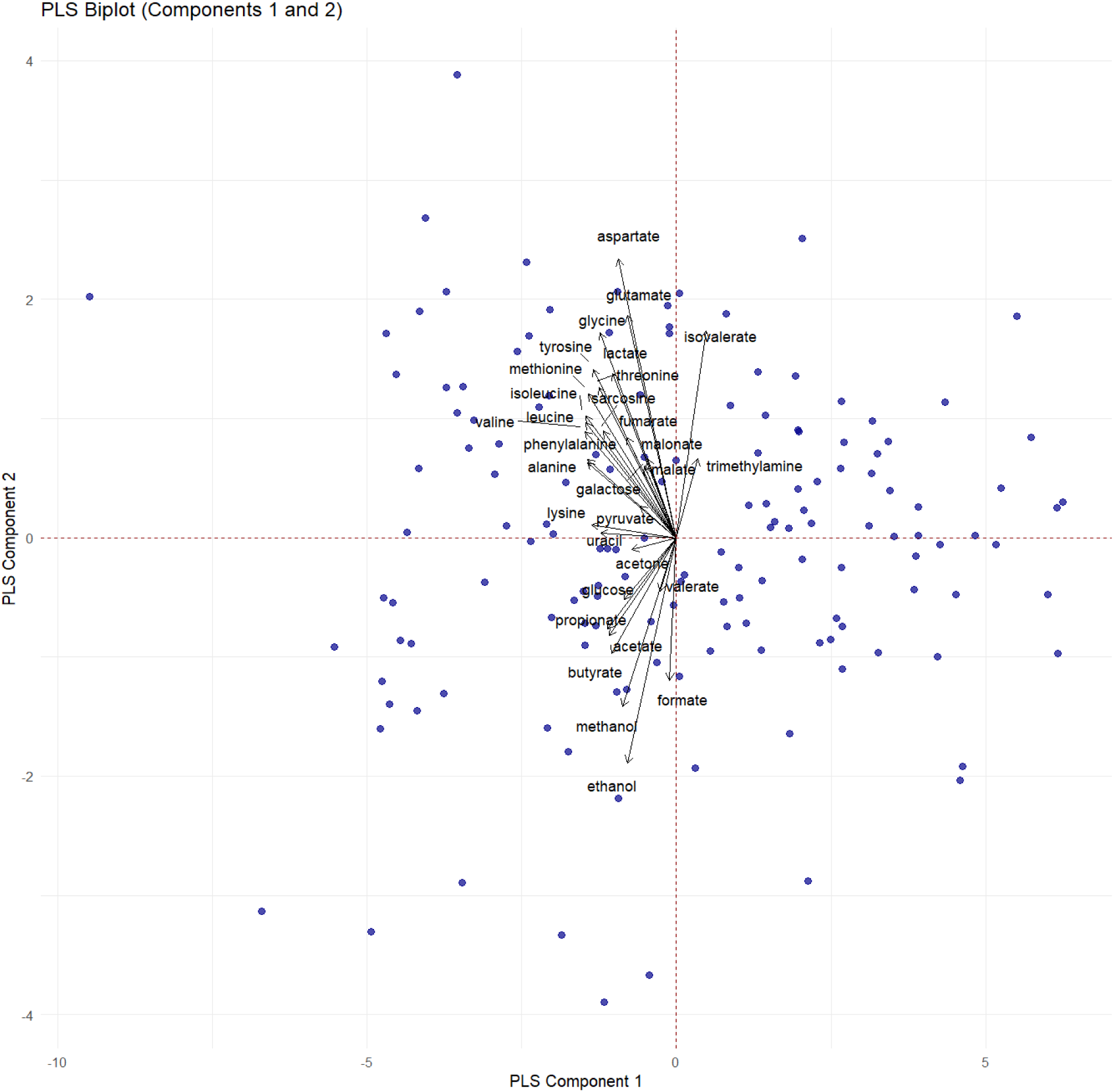
PLS2 biplot: scores and loadings of components 1 and 2, from PLS2 (BMI, HOMA-IR, WHtR)

### 3.3. Multivariate linear regression

Acetate and ethanol, which shared microbial and metabolic origins, were included in the multivariate model based on independent FDR-significant univariate associations (FDR ≤ 0.2). This combined model was used to explore their joint association with anthropometric outcomes. Among females, FFMI showed the strongest model (adj. R² = 0.09, p < 0.01), followed by WHtR (adj. R² = 0.08, p-value =0.019) and BMI (adj. R² = 0.07, p-value =0.029) (**Figure 2b**). Other anthropometric outcomes showed weak model performance (adj. R²< 0.05), suggesting limited explanatory power. In the overall population, the same model explained approximately 4.9% of the variance in BMI (adj. R² = 0.049, p-value =0.008); in females, 7% (adj. R² for acetate = 0.03, adj. R² for ethanol = 0.04). Within this model, ethanol was significantly associated with BMI (p-value =0.049), while acetate showed a statistical trend (p-value =0.060). Relative importance analysis revealed near equal contributions (ethanol 51.8%, acetate 48.2%) to the explained variance. This model was statistically significant overall (F (2, 154) = 4.997, p-value =0.008), indicating that the combination of acetate and ethanol provides a meaningful explanation of BMI variation in the full sample.

For WHtR, the same model explained 3.3% of the variance (adj. R² = 0.033, p-value =0.027; ethanol p-value =0.022, acetate p-value =0.411) with ethanol contributing 81.4% and acetate 18.6% of the explained variance. For FFMI (adj. R² = 0.030, p-value =0.039; ethanol p-value =0.109, acetate p-value =0.116), with ethanol and acetate contributing 50.8% and 49.2%, respectively. These findings suggest a small but consistent association of these metabolites, particularly ethanol, with markers of adiposity and lean mass.

### 3.4. Principal Component Analysis (PCA)

Principal component analysis of fecal metabolite profiles revealed no distinct clustering by BMI or HOMA-IR (BMI >25, HOMA-IR >2.5), suggesting limited separation based on global fecal metabolite patterns. **(Figure 3a and b**). PC1 and PC2 explained 42.6% and 11.3% variance, respectively. PC1 was driven by ethanol, acetate, glucose, and methanol, while PC2 was driven by isovalerate. BMI was weakly correlated with PC1 (r = 0.17, p-value =0.041) and uncorrelated with PC2 (r = –0.07, p-value =0.394). HOMA-IR correlated neither (r = 0.06, p-value = 0.491 for PC1 and r = - 0.07, p-value = 0.390 for PC2). Sex-stratified PCA was not conducted due to a lack of clustering in the full dataset, and stratification did not reveal an additional discriminatory pattern.

### 3.5. Partial Least Squares (PLS2) regression

*PLS2* was applied to explore the joint relationships between stool metabolites and multiple phenotypes, including BMI, HOMA-IR, and WHtR. Three-component models minimized the mean squared error of cross-validation (MSECV) for BMI, HOMA-IR, and WHtR. Model validation using root mean squared error of prediction (RMSEP) curves did not show meaningful improvement with additional components, indicating limited predictive performance. Variable Importance in Projection (VIP) scores highlighted ethanol, acetate, propionate, lysine, leucine, and isovalerate (VIP > 1) as key contributors to BMI, HOMA-IR, and WHtR **(Figure 4, Online Resource 2)**. Although amino acids and most other significant metabolites did not surpass the FDR (FDR > 0.20), all showed the same directionality in multivariate PLS2 models (VIP > 1), suggesting a consistent, albeit modest, signal worth exploration in larger cohorts.

### 3.6. Alcohol consumption and fecal metabolites

In females, ethanol concentrations were associated with alcohol intake (β = -0.05 [CI: -0.09 – -0.01], p-value = 0.022), confirming a gut-derived signal since ethanol was not used as a preservative in sample storage. Alcohol intake was also associated with glucose (β = 0.06 [CI: 0.01 – 0.10], p-value = 0.017), methanol (β = 0.05 [CI: 0.01 – 0.09], p-value = 0.019), formate (β = 0.03 [CI: 0.000 – 0.06], p-value = 0.049), and inversely with trimethylamine (TMA) (β = -0.04 [CI: -0.08 – -0.01], p-value = 0.025) in females.

In contrast, in males, alcohol intake was inversely associated with galactose (β = -0.02 [CI: - 0.05 – -0.00], p-value = 0.046) and methanol (β = -0.03 [CI: -0.07 – -0.00], p-value = 0.046) **(Online Resource 3.a)**.

### 3.7. Alcohol consumption and body composition

In females, higher alcohol consumption was positively associated with several anthropometric traits related to both visceral and subcutaneous adiposity. These included BMI (β = 0.41 [CI: 0.14–0.67, p-value = 0.002), WHtR (β = 0.64 [CI: 0.22–1.07], p-value = 0.003), and FFMI (β = 0.14 [CI: 0.04–0.25], p-value = 0.007).

Conversely, alcohol consumption was negatively relationship between a associated with the muscle-to-fat ratio in females (β =-0.05 [CI: -0.09 – -0.01], p-value = 0.022), potentially reflecting a shift toward higher fat relative to muscle mass. Also, body fat percentage showed a positive association with alcohol intake in females (β = 0.46 [CI: 0.07–0.86], p-value = 0.023), supporting increased relative adiposity rather than a ratio-driven effect.

No significant association was found between alcohol intake and HOMA-IR in either sex (Online Resource 3.b).

### 3.8. Energy and energy-related mediators

Several fecal metabolites were associated with BMR and TEE. After FDR correction, acetate, butyrate, glucose (FDR≤ 0.05, adj. R² = 0.14, 0.13, and 0.12), and methanol (FDR = 0.025, adj. R² = 0.11) remained significantly associated with TEE in females **(Figure 5)**. No significant associations were observed in males. Moreover, no metabolites remained significant for BMR after FDR correction **(Online Resource 4. a and b).**

**Figure 5.**
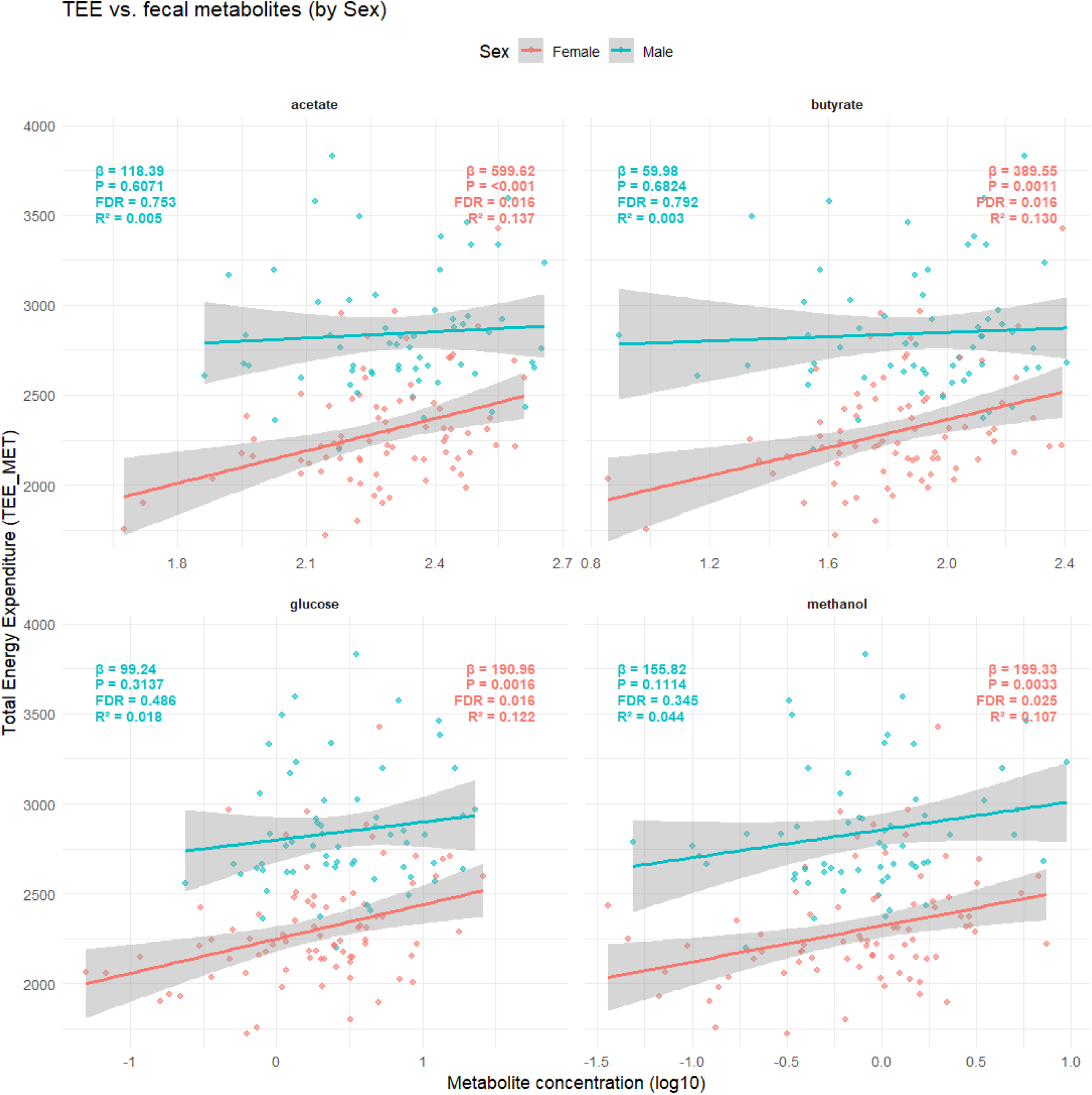
Total energy expenditure (TEE) vs. fecal metabolites using sex-stratified multiple linear regression analysis with Significant FDR:

## 4. Discussion

Our study highlights that specific gut microbiota-derived metabolites are significantly associated with key body composition traits, energy expenditure, and alcohol intake in adolescents, particularly in females. Acetate, as an SCFA and an important substrate for hepatic lipogenesis and gluconeogenesis, as well as a signaling metabolite implicated in muscle biology, has been previously linked to adiposity and metabolic regulation (Tremaroli & Bäckhed, 2012; Yang et al., 2024). In line with this literature, our sex-stratified analyses revealed that fecal acetate was positively associated with BMI in females, and, together with ethanol, contributed to multivariate models for FFMI and WHtR.

Multivariate models, including acetate and ethanol, captured lean and central adiposity, particularly FFMI and WHtR, with greater specificity than BMI. This observation is consistent with evidence that metabolite profiles can reflect body-composition phenotypes beyond total mass(Baars et al., 2018; Fernández-Verdejo et al., 2024; Vallianou et al., 2019) and with reviews positioning microbiota-derived metabolites as central regulators in metabolic disorders (Borrego-Ruiz & Borrego, 2025). FFMI, by correcting for height and distinguishing between fat and fat-free compartments, offers an advantage over BMI (Chambers et al., 2019), while WHtR is a robust indicator of visceral adiposity(Ashwell & Gibson, 2016). Both measures showed stronger associations with fecal metabolites than BMI in our models, aligning with prior findings that central and lean mass compartments are more sensitive to metabolic disorders, and are supported by RCT meta-analytic evidence demonstrating reductions in body weight, waist circumference, and fat mass (Chahibakhsh et al., 2024).

Several other metabolites also showed nominal associations with anthropometrics in females, although they did not survive FDR correction. For example, amino acids leucine, lysine, and alanine were positively related to BMI, and lysine was further associated with FFMI (Newgard et al., 2009). These findings align with prior reports that branched-chain and essential amino acids are linked to obesity and IR through their role in lipid and energy metabolism (Wang et al., 2011).

Also, propionate was positively associated with BMI, and this SCFA is noteworthy given its established insulin-sensitizing properties in human intervention studies (Chambers et al., 2018). Moreover, formate, a metabolite linked to acetate metabolism and one-carbon pathways, also demonstrated associations with anthropometry, consistent with previous reports linking formate to metabolic flux and adiposity (Mardinoglu et al., 2015; Pietzke et al., 2020). Although these associations did not meet FDR thresholds, the fact that they recurred across univariate and multivariate models suggests biological plausibility but also highlights limited statistical power in exploratory metabolomics cohorts, where small sample sizes increase the risk of false negatives, and indicates that larger cohorts may be required to validate these trends (Baars et al., 2018; Zierer et al., 2018). This limitation is consistent with recent pediatric metabolomics studies, reporting SCFA and amino acid associations that attenuate after correction for multiple testing (Li et al., 2025; Wu et al., 2023). In contrast, a recent longitudinal COPSAC birth-cohort analysis with repeated early-life stool sampling reported no consistent associations between early-life gut microbiota composition and later childhood BMI development or DXA-derived body composition. Together with our findings, this suggests an age-dependent model of gut–metabolism relationships, in which microbial contributions to adiposity become more detectable later in development, potentially reflecting increasing influence of hormonal maturation and lifestyle-related exposures (Poulsen et al., 2025).

PCA and PLS2 analyses provided additional insights. PCA did not reveal strong clustering by BMI or HOMA-IR, underscoring the complexity of fecal metabolite patterns and their modest ability to discriminate metabolic phenotypes in this age group. PC1 was largely driven by ethanol, acetate, glucose, and methanol, while PC2 was dominated by isovalerate. Consistent with this, PLS2 regression highlighted ethanol, acetate, propionate, lysine, leucine, and isovalerate as key contributors (VIP>1) across BMI, WHtR, and HOMA-IR models. Importantly, isovalerate, a branched-chain fatty acid produced by microbial fermentation of leucine, has been previously linked to obesity and impaired glucose regulation(Le Chatelier et al., 2013; Pedersen et al., 2016). While its associations in our cohort were modest and not FDR significant, the consistency across PCA and PLS2 suggests potential relevance of isovalerate as a potential indicator of protein fermentation and metabolic perturbation (Wu et al., 2023).

Beyond intrinsic metabolite–anthropometric associations, alcohol intake, examined as a post hoc exposure, was associated with fecal metabolite profiles and body composition. In females, alcohol consumption was positively associated with fecal ethanol, methanol, glucose, and formate, consistent with established alcohol fermentation and metabolism pathways (Baraona & Lieber, 1979; Cederbaum, 2012; Engen et al., 2015). At the same time, alcohol intake was inversely associated with TMA, a finding that contrasts with alcohol-associated liver disease, where TMA is typically elevated. One possible explanation is that alcohol suppresses choline-utilizing, TMA-producing microbes, leading to reduced TMA levels in moderate drinkers, particularly in females. In contrast, in alcohol-related liver disease, pathological dysbiosis elevates TMA levels due to impaired microbial and hepatic regulation (Chen et al., 2016; Hoyles et al., 2018; Romano et al., 2015). These findings suggest that alcohol’s effects on TMA may be context-dependent, differing between moderate consumption in healthy individuals and disease states. In males, alcohol consumption showed negative associations with galactose and methanol, which highlights potential sex-specific differences in microbial fermentation dynamics (Furuhashi & Hotamisligil, 2008). Recent microbiome studies confirm such sex-specific metabolic interactions, particularly in relation to metabolic syndrome and obesity(Garcia-Fernandez et al., 2024).

These microbial changes coincided with anthropometric effects. In females, higher alcohol intake was associated with increased BMI and WHtR, while the muscle-to-fat ratio decreased, suggesting a shift in body composition toward higher fat mass relative to lean tissue. Such findings are consistent with epidemiological evidence linking alcohol intake to central adiposity and altered body composition (Traversy & Chaput, 2015). Notably, no associations were detected between alcohol intake and HOMA-IR, underscoring the complexity of alcohol’s metabolic effects and the potential for phenotype-specific responses.

While BMR and TEE were estimated from anthropometric inputs, all associations were evaluated using FDR correction, supporting the robustness of the observed relationships despite potential collinearity. Acetate, butyrate, glucose, and methanol were positively associated with TEE in females, suggesting a contribution of gut microbial metabolites to host energy homeostasis that may modulate metabolic efficiency. Acetate and butyrate, the two main SCFAs, are known to participate in mitochondrial energy pathways and muscle metabolism, which may reflect interindividual variability in energy expenditure through effects on diet-driven energy turnover and substrate oxidation (Benítez-Páez et al., 2021; Corbin et al., 2023; Tremaroli & Bäckhed, 2012; Yang et al., 2024). These findings extend existing evidence by implicating gut-derived metabolites as an additional regulatory layer and are consistent with reported sex-related differences in SCFA–energy metabolism coupling, potentially reflecting hormonal or mitochondrial influences (Bellissimo et al., 2022; Kadyan et al., 2023)

In contrast, classical energy intermediates such as lactate and pyruvate were not associated with energy expenditure, which may reflect compartmentalization of systemic versus microbial metabolism or limitations of fecal metabolomics in capturing central energy fluxes. By comparison, microbiota-derived metabolites, including acetate, butyrate, glucose, and methanol, were consistently linked to TEE, emphasizing that fecal metabolomics captures microbial contributions to host metabolism that extend beyond central host-derived pathways (Baars et al., 2018).

Notably, such associations were evident even in participants with normal BMI, highlighting the limitation of traditional anthropometric measures in identifying subtle metabolic risk. Integrating metabolomic data with compositional indicators, such as FFMI or WHtR, may enable more accurate detection of early metabolic dysfunction (Borrego-Ruiz & Borrego, 2025; Chambers et al., 2019). These findings strengthen the biological plausibility of a link between gut-derived SCFAs and host energy metabolism, particularly in females. These findings reinforce the idea that microbial metabolites serve as biomarkers of fat mass, fat mass distribution, and metabolic risk, which can be intensified by lifestyle and personal patterns, including alcohol consumption. Females appear more influenced by fecal metabolites (Zmora et al., 2019), potentially due to sex-related differences in microbiota–host interactions, and may also be more prone to insulin resistance and systemic inflammation (Garcia-Fernandez et al., 2024; Olalekan et al., 2024).

This study has several strengths. We applied multiple complementary body composition measures beyond BMI, particularly FFMI and WHtR, which allowed a more nuanced characterization of adiposity and lean mass distribution. In addition, sex-stratified analyses and the combined use of univariate, multivariate, and projection-based methods, such as PCA and PLS2, strengthened the robustness and interpretability of the findings. Finally, the analysis was conducted in a well-characterized adolescent cohort with detailed metabolic and lifestyle data. Several limitations should also be considered. The exploratory cross-sectional design precludes causal inference, and the observed associations can not establish directionality. The use of NMR spectroscopy, while well-suited for absolute quantification, has lower sensitivity than mass spectrometry, and data on stool transit time and the virome were not available. In some studies, it is indicated that there are other factors influencing the concentration of the metabolites affecting the SCFAs in the stool samples, such as the stool consistency and intestinal transit time, which strongly influence SCFA production and fecal concentrations, with slower transit times leading to increased proteolysis and altered SCFA profiles (Vandeputte et al., 2016). These factors likely contribute to inter-individual variability in our metabolite associations, and they were not evaluated in this study.

Future longitudinal multi-omics studies integrating metabolomics, microbiome composition, dietary intake, and hormonal factors, together with mechanistic approaches, are needed to clarify causality and the interplay between microbes, diet, and host metabolism.

## Supporting information

Alcohol consumption questions

## Data Availability

The data supporting the findings of this study are not publicly available due to ethical and legal restrictions related to participant privacy, but can be made available upon reasonable request, subject to approval by the corresponding author.

## Acknowledgements

We extend our sincere gratitude to all participants of the Copenhagen Prospective Studies on Asthma in Childhood (COPSAC) for their essential contributions to this research. We also gratefully acknowledge the financial support provided by the Lundbeck Foundation, the Danish Ministry of Health, the Danish Council for Strategic Research, and the Capital Region Research Foundation.

## Author Contributions

MAR has designed the experiment. PE and MAR have supervised, and NCH analyzed the data and wrote the main manuscript text and prepared all figures and tables. All the authors have contributed to the scientific discussion and approved the final manuscript.

## Ethical approval

The study was approved by the Ethics Committee of the Capital Region of Denmark (H-16040846) and the Danish Data Protection Agency (2015-41-3696). Written informed consent was obtained from all participants (or their legal guardians, where applicable).

