## Supplemental tables and figures for "The Association Between Gut-Derived Metabolites, Body Composition, and Energy Expenditures in Adolescents: A Sex-Stratified Analysis of the COPSAC2000 Study": Supplementary material-Illustration Fecal metabolomics 09.02.2026.docx

**Online Resource 1. Supplementary Table S1.** Descriptive fecal metabolite composition and concentration characteristics of the participants. The first number under each column is the median, and the numbers in the parentheses are the 25th percentile (Q1) and the 75th percentile (Q3), respectively, showing the Inter-Quartile Range (IQR).

| **Fecal Metabolite millimolar (mM)** | **Female^1^** | **Male^1^** | **p_value^2^** | **FDR^3^** |
| --- | --- | --- | --- | --- |
| **acetate** | 195.2 (152.1–266.84) | 209.05 (159.12–289.76) | 0.523 | 0.795 |
| **acetone** | 0.31 (0.15–0.68) | 0.47 (0.28–0.76) | 0.038**^*^** | 0.417 |
| **alanine** | 8.93 (6.59–12.29) | 10.16 (6.85–13.82) | 0.244 | 0.66 |
| **aspartate** | 1.95 (1.37–2.83) | 1.86 (1.36–2.66) | 0.947 | 0.947 |
| **butyrate** | 74.12 (49.16–106.31) | 83.48 (47.47–132.78) | 0.244 | 0.66 |
| **ethanol** | 0.87 (0.39–1.78) | 0.97 (0.38–2.16) | 0.755 | 0.933 |
| **formate** | 0.14 (0.09–0.2) | 0.14 (0.1–0.18) | 0.609 | 0.847 |
| **fumarate** | 0.2 (0.11–0.31) | 0.25 (0.13–0.4) | 0.029**^*^** | 0.417 |
| **galactose** | 0.39 (0.25–0.54) | 0.43 (0.25–0.67) | 0.232 | 0.66 |
| **glucose** | 2.06 (0.94–4.07) | 2.39 (1.18–6.91) | 0.126 | 0.66 |
| **glutamate** | 5.39 (3.96–6.99) | 5.35 (3.79–7) | 0.847 | 0.947 |
| **glycerol** | 2.33 (1.33–3.85) | 2.05 (1.35–3.86) | 0.697 | 0.93 |
| **glycine** | 3.18 (2.63–4.67) | 3.55 (2.39–4.08) | 0.933 | 0.947 |
| **isoleucine** | 16.23 (11.62–22.11) | 17.34 (11.53–24.41) | 0.398 | 0.75 |
| **isovalerate** | 16.01 (8.22–27.55) | 13.03 (5.97–24.08) | 0.25 | 0.66 |
| **lactate** | 3.24 (2.37–4.34) | 2.88 (2.33–4.45) | 0.758 | 0.933 |
| **leucine** | 9.74 (6.65–12.69) | 10.42 (6.8–14.07) | 0.385 | 0.75 |
| **lysine** | 7.45 (5.36–11.05) | 9.22 (6.24–13) | 0.065 | 0.417 |
| **malate** | 0.62 (0.34–1.05) | 0.71 (0.45–1.22) | 0.233 | 0.66 |
| **malonate** | 0.33 (0.21–0.64) | 0.46 (0.24–0.84) | 0.054 | 0.417 |
| **methanol** | 0.99 (0.42–1.74) | 0.97 (0.44–1.46) | 0.547 | 0.795 |
| **methionine** | 3.56 (2.5–5.63) | 3.79 (2.46–5.87) | 0.528 | 0.795 |
| **phenylalanine** | 3.15 (2.35–4.14) | 3.44 (2.39–4.74) | 0.244 | 0.66 |
| **propionate** | 87.11 (61.4–111.49) | 91.87 (62.17–123.28) | 0.49 | 0.795 |
| **pyruvate** | 1.44 (0.83–3.1) | 1.63 (0.91–4.22) | 0.443 | 0.787 |
| **sarcosine** | 0.21 (0.13–0.29) | 0.2 (0.12–0.29) | 0.891 | 0.947 |
| **threonine** | 1.77 (1.12–2.6) | 1.88 (1.2–2.57) | 0.831 | 0.947 |
| **trimethylamine** | 0.48 (0.14–0.77) | 0.58 (0.33–0.87) | 0.057 | 0.417 |
| **tyrosine** | 2.32 (1.65–3.36) | 2.69 (1.78–3.7) | 0.373 | 0.75 |
| **uracil** | 0.94 (0.74–1.19) | 1.06 (0.75–1.27) | 0.268 | 0.66 |
| **valerate** | 16.36 (10.62–20.46) | 15.84 (8.94–22.53) | 0.911 | 0.947 |
| **valine** | 11.76 (8.01–16.55) | 12.69 (8.13–18.48) | 0.388 | 0.75 |

**^1^Median (IQR)**

**^2^Wilcoxon rank sum test**

**^3^Benjamini-Hochberg method (FDR) correction**

**^*^ Significant p_value < 0.05**

**Online Resource 2. Supplementary Figure S1.** Variable Importance in Projection (VIP) scores from partial least squares (PLS) regression. Metabolites with VIP scores > 1 are highlighted as major contributors to the multivariate models.

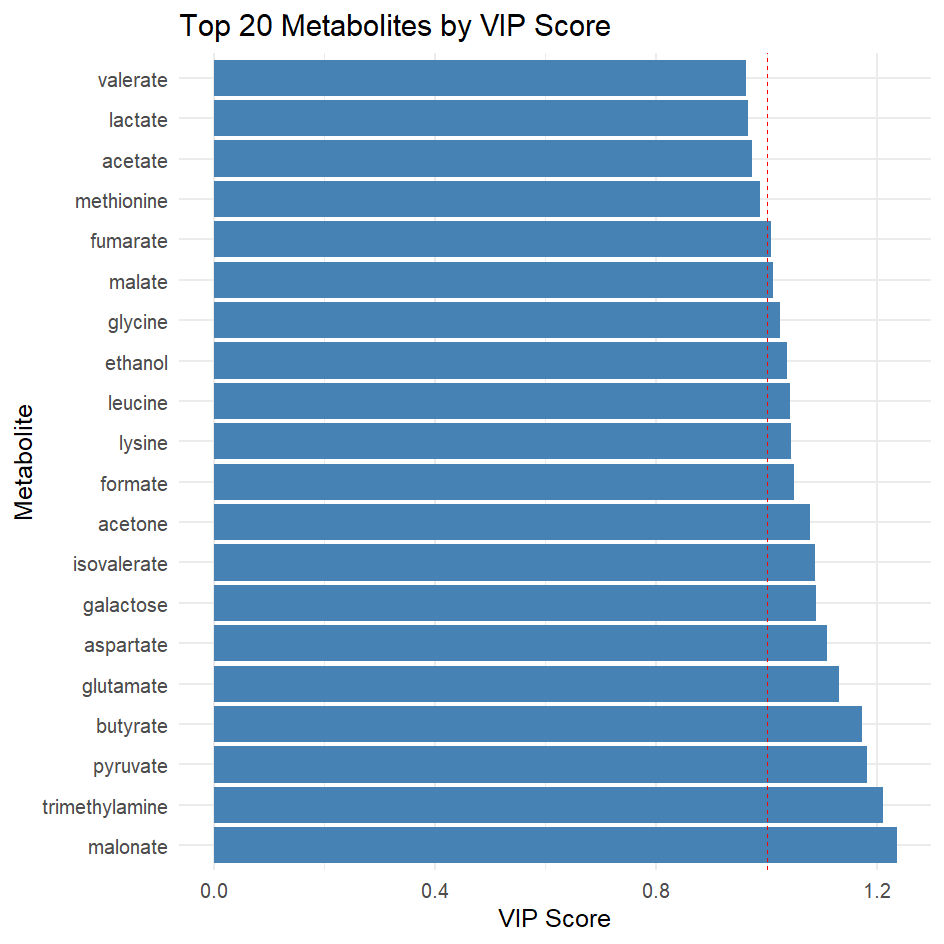

**Online Resource 3. Supplementary Figure S2.** Associations between alcohol consumption, fecal metabolites, and body anthropometric traits. Panels show sex-stratified linear regression analyses of alcohol intake with a. Fecal metabolites, and b. Body composition measures and HOMA-IR.

a.
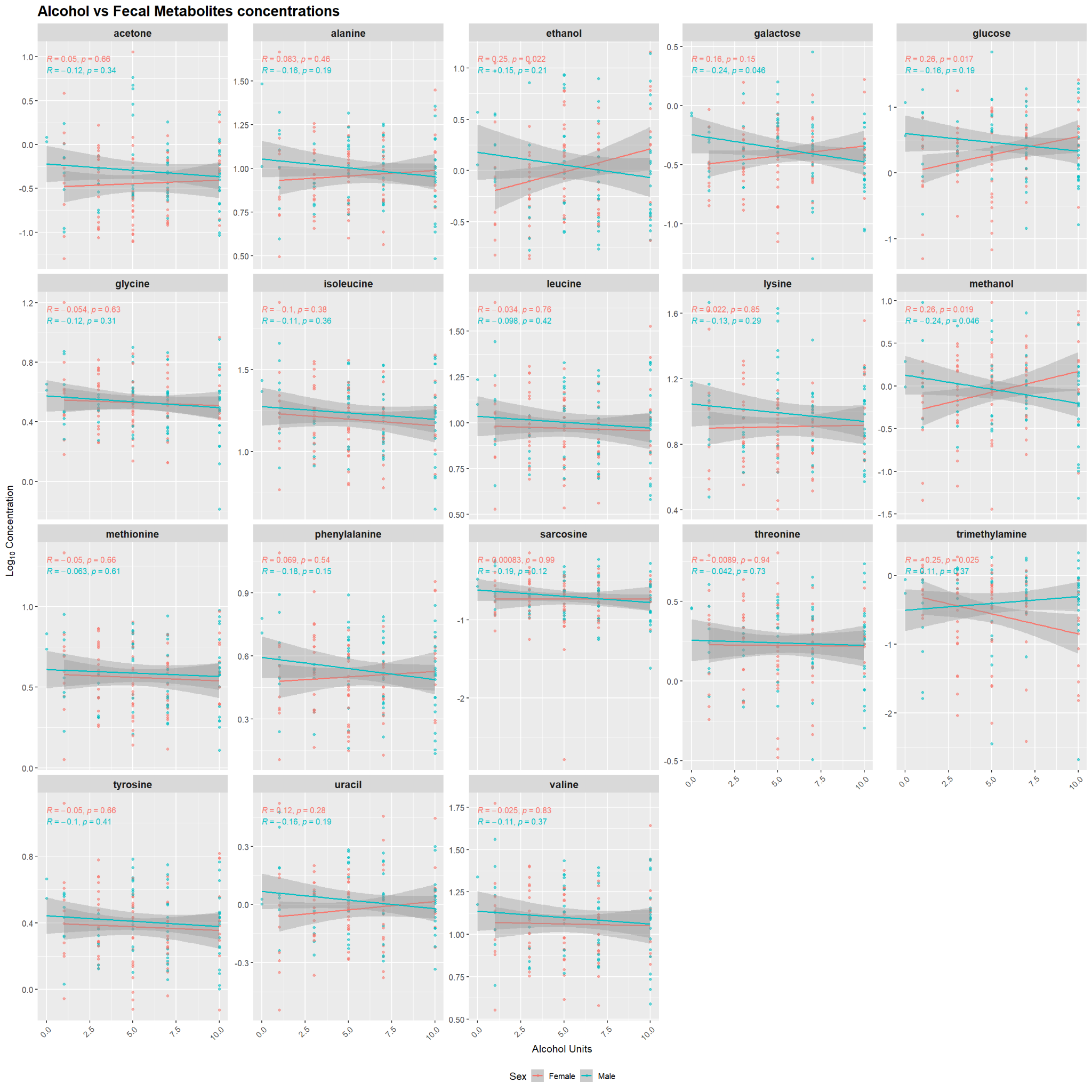

b.

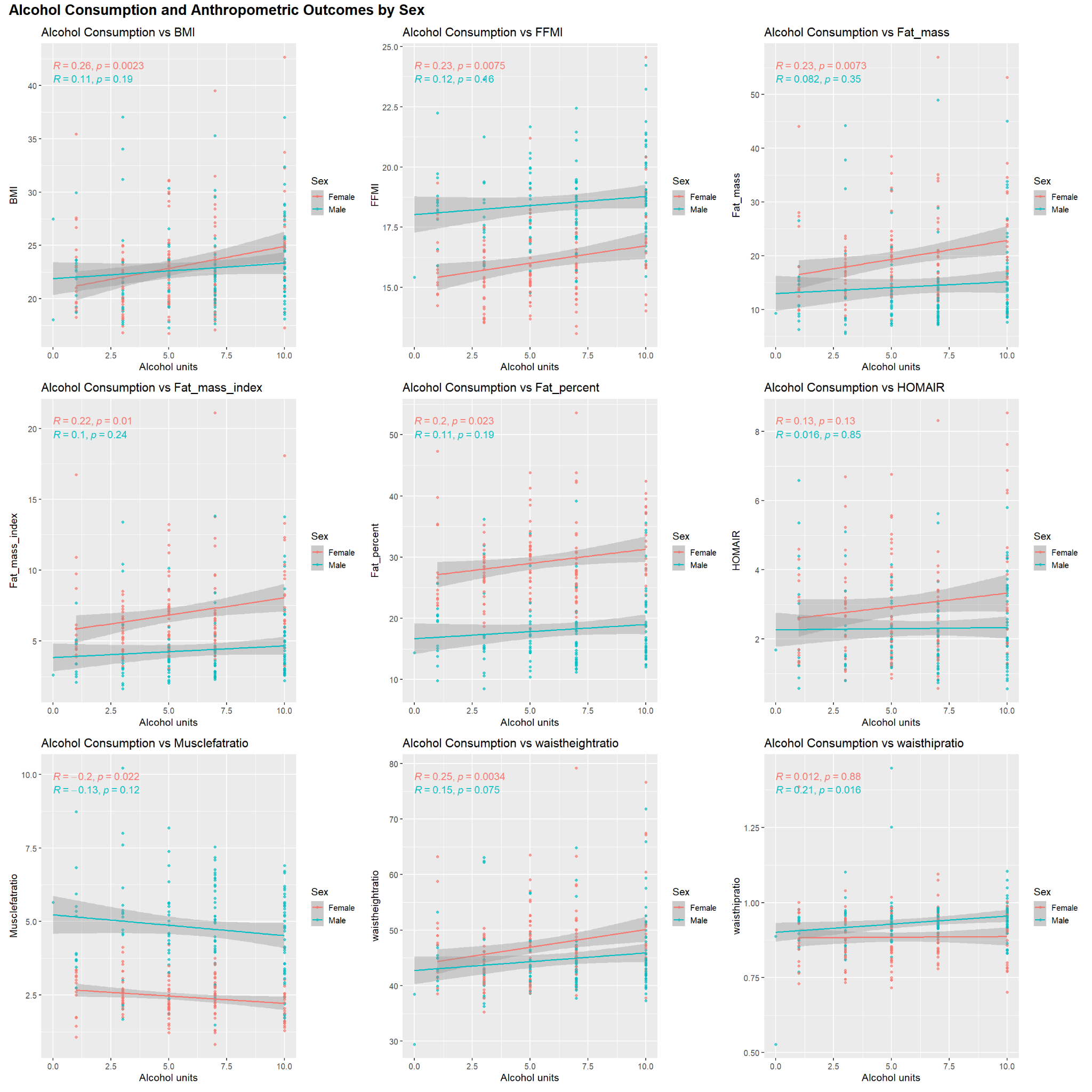

**Online Resource 4. Supplementary Figure S3.** Sex-stratified associations between fecal metabolites and energy expenditure; (a) Basal metabolic rate (BMR) and fecal metabolites, and (b) Total energy expenditure (TEE) and fecal metabolites.

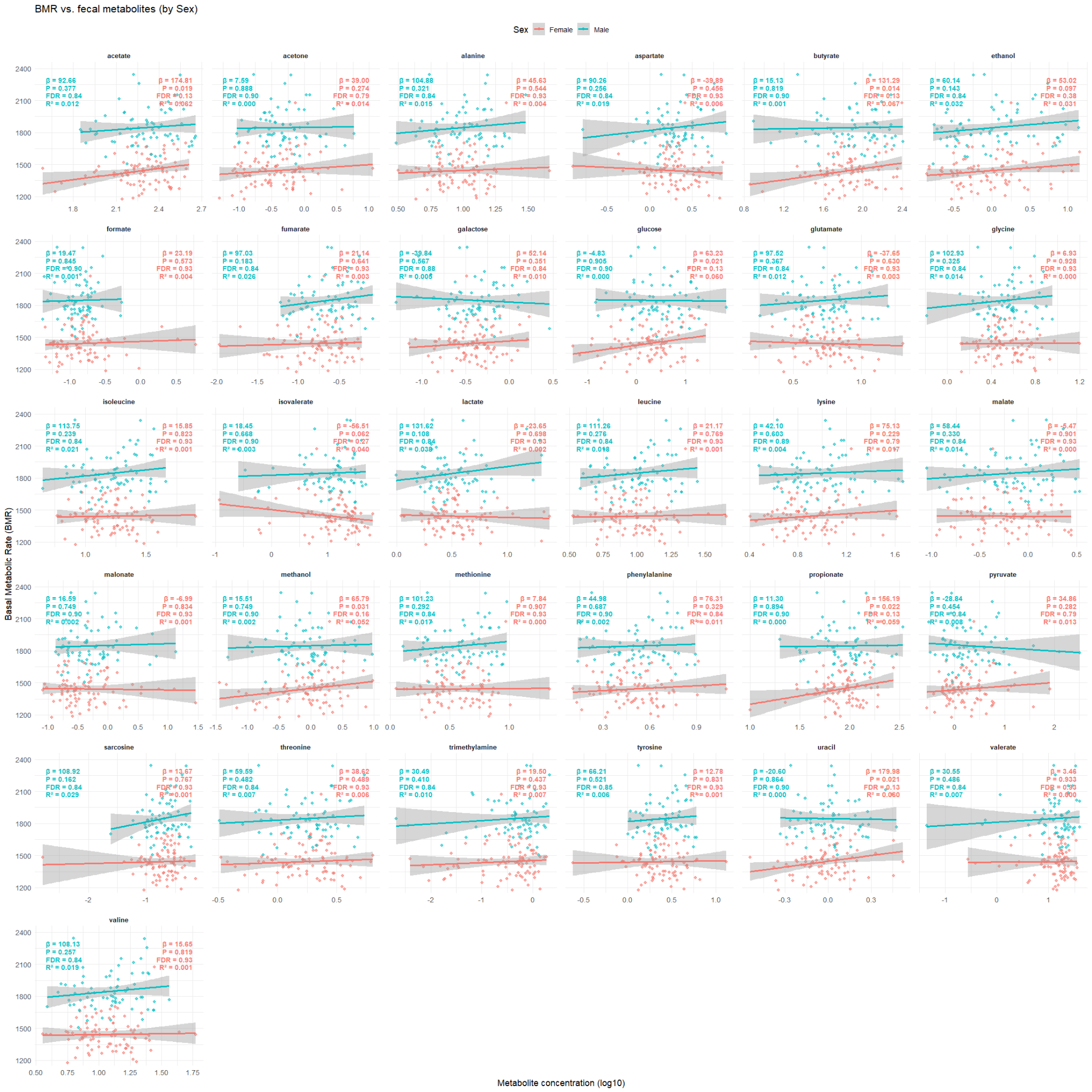

**b.**

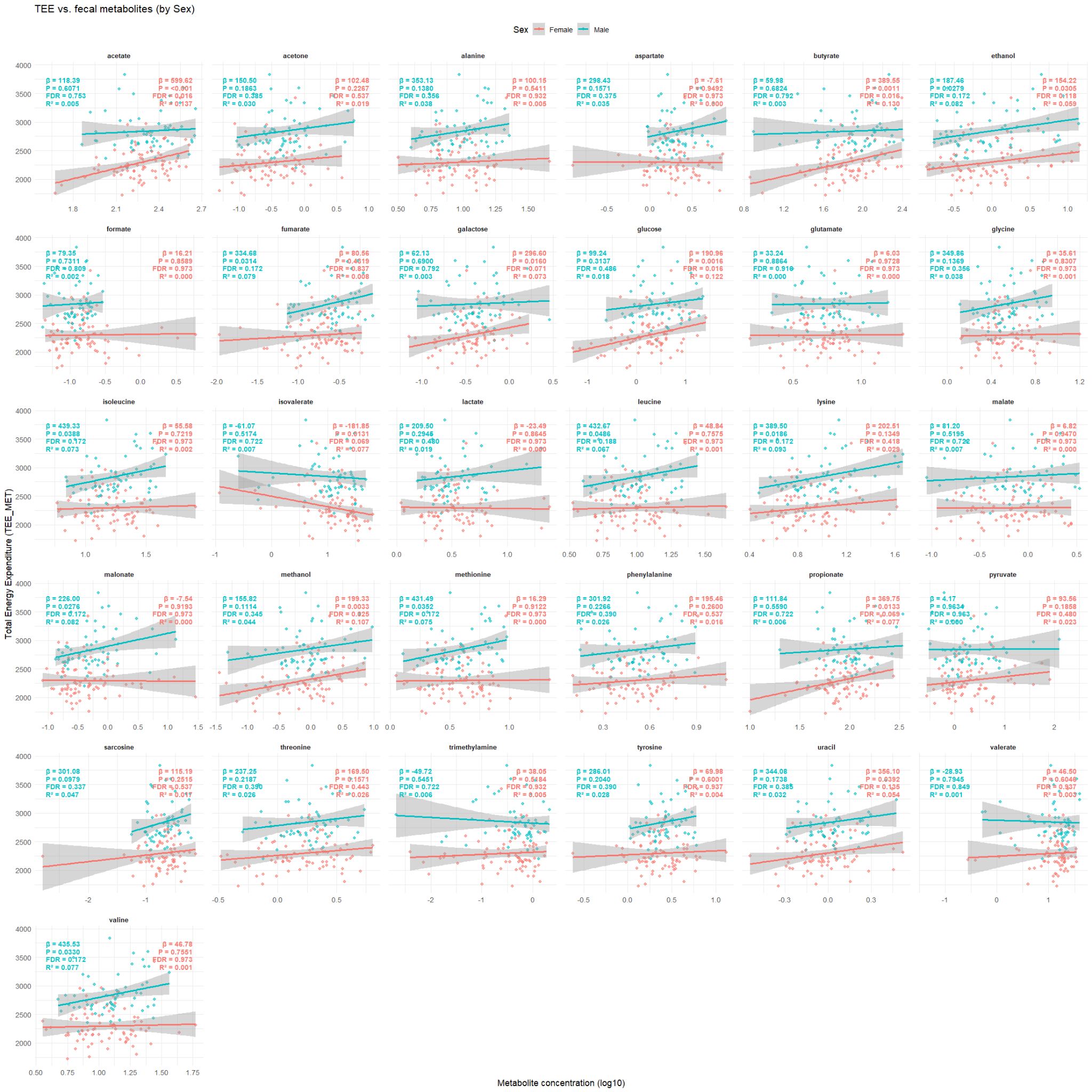
