## Supplementary material for "The Association Between Gut-Derived Metabolites, Body Composition, and Energy Expenditures in Adolescents: A Sex-Stratified Analysis of the COPSAC2000 Study": Alcohol consumption questions: supplementary alcohol consumption questions.docx

**Supplementary Methods: Assessment of Alcohol Consumption**

Alcohol consumption was assessed at age 18 using a self-reported questionnaire designed to capture key aspects of drinking behavior.

Participants answered five items addressing lifetime exposure, age at initiation, frequency, quantity, and binge drinking patterns

The following questions:

1. Have you ever consumed more than one unit of alcohol on a single occasion?

(Yes/No)

2. How old were you the first time you drank more than one unit of alcohol?

(Years)

3. How often do you drink something that contains alcohol?

(Frequent)

4. How many units of alcohol do you typically consume when you drink?

(From 1 to greater than 10)

5. How often do you drink five or more units of alcohol on a single occasion?

(Never, Weekly, Less One Monthly, Monthly)

Alcohol intake was quantified based on self-reported responses. One unit of alcohol was defined according to the Danish Health Authority guidelines as containing 12 g of pure alcohol.
